# Automatic detection of simulated artifacts on T1w magnetic resonance images: comparing performance of different QC strategies

**DOI:** 10.1101/2025.10.31.25339144

**Authors:** Janine Hendriks, Michelle G. Jansen, Richard Joules, Óscar Peña-Nogales, Paulo R. Rodrigues, Frederik Barkhof, Anouk Schrantee, Henk J.M.M. Mutsaerts, the Alzheimer’s Disease Neuroimaging Initiative

## Abstract

The reliability of MRI-derived measures critically depends on image quality. Poor-quality scans can obscure anatomical detail and compromise the accuracy of automated image analysis, underscoring the need for robust quality control (QC) procedures. Automated QC offers scalability for large neuroimaging datasets, yet the comparative performance of different approaches for detecting specific artifact types remains poorly understood.

We systematically compared rule-based (RB), classical machine learning (ML), and deep learning (DL) QC algorithms using 1,000 high-quality T1w scans. Four artifact types, blurring, ghosting, motion, and noise were synthetically introduced across ten severity levels using TorchIO, yielding 40,000 degraded images. Visual QC of a subset confirmed strong inter-rater reliability (Krippendorff’s α=0.82, mean Spearman’s ρ=0.87). RB and ML models used 62 image quality metrics (IQMs) from MRIQC, whereas DL models were trained directly on minimally preprocessed images. Models were trained with participant-level five-fold cross-validation and tested on an independent dataset.

DL models achieved the highest overall performance across artifact types (Youden’s Index=0.83–0.97). RB and ML performed comparably at high artifact severities (YI≥0.75) but showed limited sensitivity to subtle ghosting and noise (YI≤0.15). Feature analysis indicated that RB relied primarily on normative metrics, whereas ML flexibly adapted feature use by artifact type and severity.

These findings highlight DL’s superior generalizability for detecting subtle artifacts and provide practical guidance for selecting QC strategies in large-scale neuroimaging pipelines, where reliable QC is essential for maintaining statistical power and reproducibility.

## 1. Introduction

Magnetic Resonance Imaging (MRI) is a powerful, non-invasive diagnostic tool that provides detailed images of brain anatomy, making it invaluable for investigating neurological conditions in both research and clinical settings. T1-weighted images are frequently used to derive conventional measures of brain health, such as gray matter volume or cortical thickness. However, the reliability of MRI derivatives is heavily dependent on the quality of the acquired MRI scans. Poor image quality not only makes it challenging for radiologists to discern relevant anatomical detail, but may also compromise the accuracy and precision of automated image analysis algorithms (Alexander-Bloch et al., 2016; Perrone et al., 2015; Power et al., 2012; Reuter et al., 2015). Therefore, robust quality control (QC) measures are essential to identify and appropriately manage poor-quality images that could negatively impact statistical power in research studies. Beyond guiding exclusion decisions, QC metrics can also be used to modulate confidence in results, serving as covariates or weights in subsequent analyses (Birn, 2023; Taylor et al., 2023).

Traditional QC methods typically involve visual inspection by radiographers, radiologists, or researchers, which is both time-consuming and subjective (Scheltens et al., 1995; Williams et al., 2023). In contrast, automated QC could offer significant advantages, such as improved reproducibility and standardization by eliminating human bias, and increased efficiency because of computer speed and parallelization potential. These advantages make automated QC a feasible solution for large research studies or potentially even real-time feedback on the scanner in the future. Despite these benefits, the implementation of automated QC has yet to be standardized across institutions, in part due to the diversity of research protocols, scanner hardware, and study aims. Research groups often develop their own QC algorithms to assess image quality, particularly focusing on artifact detection before further post-processing (Alfaro-Almagro et al., 2018; Esteban et al., 2017; Hendriks et al., 2024). Some of these algorithms perform the assessment fully automated (automatic QC), while others still require human intervention at key points (semi-automatic QC). As the field advances, establishing consensus on systematic QC approaches is essential for enhancing the reliability of studies and improving comparability across different research centers and clinical settings.

(Semi-)automatic QC algorithms can be categorized into three categories: rule-based (RB), classical machine learning (ML), and deep learning (DL) approaches (Hendriks et al., 2024). RB approaches establish thresholds for predefined image quality features, such as signal-to-noise ratio (SNR), to flag scans that fall outside these thresholds (Kim et al., 2019). ML algorithms rely on similar predefined features but involve the training of classifiers to differentiate image quality levels (Esteban et al., 2017). DL algorithms differ from RB and ML as DL does not rely on predefined quality features but instead uses all available image information to classify the images (Bottani et al., 2022; Fantini et al., 2021; Keshavan et al., 2019).

Algorithms from each of these categories have demonstrated high accuracy within (mostly) single-cohort studies, which makes it challenging to compare QC studies due to variations in MRI scanner hardware, MRI protocols, and patient populations (Kruggel et al., 2010). The variability introduced by these factors can lead to discrepancies in image characteristics that are not accounted for by the algorithms trained on a specific dataset. Moreover, the relative performance of these methods is difficult to quantify, as reported accuracy is often contingent on the specific context of the studies and local practices, rather than on universally applicable quality criteria (Hendriks et al., 2024). The lack of standardized benchmarking datasets and evaluation metrics further complicates the comparison of different QC methods. For example, some studies may emphasize sensitivity to certain artifacts over others, such as blurring, noise, head motion or ghosting, which can bias the reported performance metrics (Krupa and Bekiesinska-Figatowska, 2015).

The present study compared the performance of automatic QC algorithms in a controlled setting using synthetic artifacts within a single large dataset, to better understand their strengths and limitations, ultimately supporting the development and implementation of automated MRI QC in research and clinical practice. We 1) evaluate whether different severity levels of these synthetic artifacts can be detected visually by human readers, 2) compare the artifact detection performance between RB, ML, and DL, and 3) investigate whether IQM feature importance differs between RB and ML.

## 2. Methods

### 2.1. Data inclusion

Data for this study were drawn from the Alzheimer’s Disease Neuroimaging Initiative (ADNI) dataset, a multisite study of older adults with normal cognition, mild cognitive impairment, or Alzheimer’s disease dementia (Petersen et al., 2010).

We selected high quality 3D T1-weighted MRI scans that are free of apparent artifacts, using visual QC scores from three independent groups. From the full pool of 17,141 3D T1w MRI scans (Jack et al., 2024, downloaded August 2024), 1,806 3T images had three visual QC scores available for the unprocessed T1w scans, one score from each of the three independent sources: the Dementia Research Centre of University College London (UCL), the NeuroImaging & Surgical Technologies (NIST) Lab of McGill University, and the Public Open Neuroimaging Data Reduction for Artificial Intelligence (PondrAI) QC. The UCL laboratory performed visual QC using a 3-level scale: 1) “ok”, 2) “borderline”, and 3) “fail”. Their assessment focused on identifying the presence of artifacts such as blurring, ghosting, motion, noise, and ringing (Manning et al., 2017). The McGill laboratory involved 12 raters who performed visual QC using QRater with a 3-level scale: 1) “ok”, 2) “warning”, and 3) “fail”. The QRater visual QC protocol outlines criteria based on coverage, intensity uniformity, motion, and noise (Fernandez-Lozano et al., 2022). PondrAI QC is a public repository containing visual QC of a variety of public datasets. The images were rated by a single rater on a 6-level scale, with 1 indicating good image quality and 6 indicating bad image quality, based on contrast, coverage, noise, and ringing (Costantino and Devenyi, 2023).

We rescaled all ratings to a 3-point scale (1-3), with 1 indicating good image quality and 3 indicating bad image quality, and summed them to a composite QC score (3-9). A total of 1,462 scans received a composite QC score of 3. Using a fixed random seed, we pseudorandomly selected a subset of 1,000 scans for subsequent analysis (for acquisition details, see Table 1). These 1,000 MRI scans included data from 694 unique participants; 445 participants had only one session, 200 had two sessions, 41 had three sessions, and 8 had four sessions.

**Table 1.**
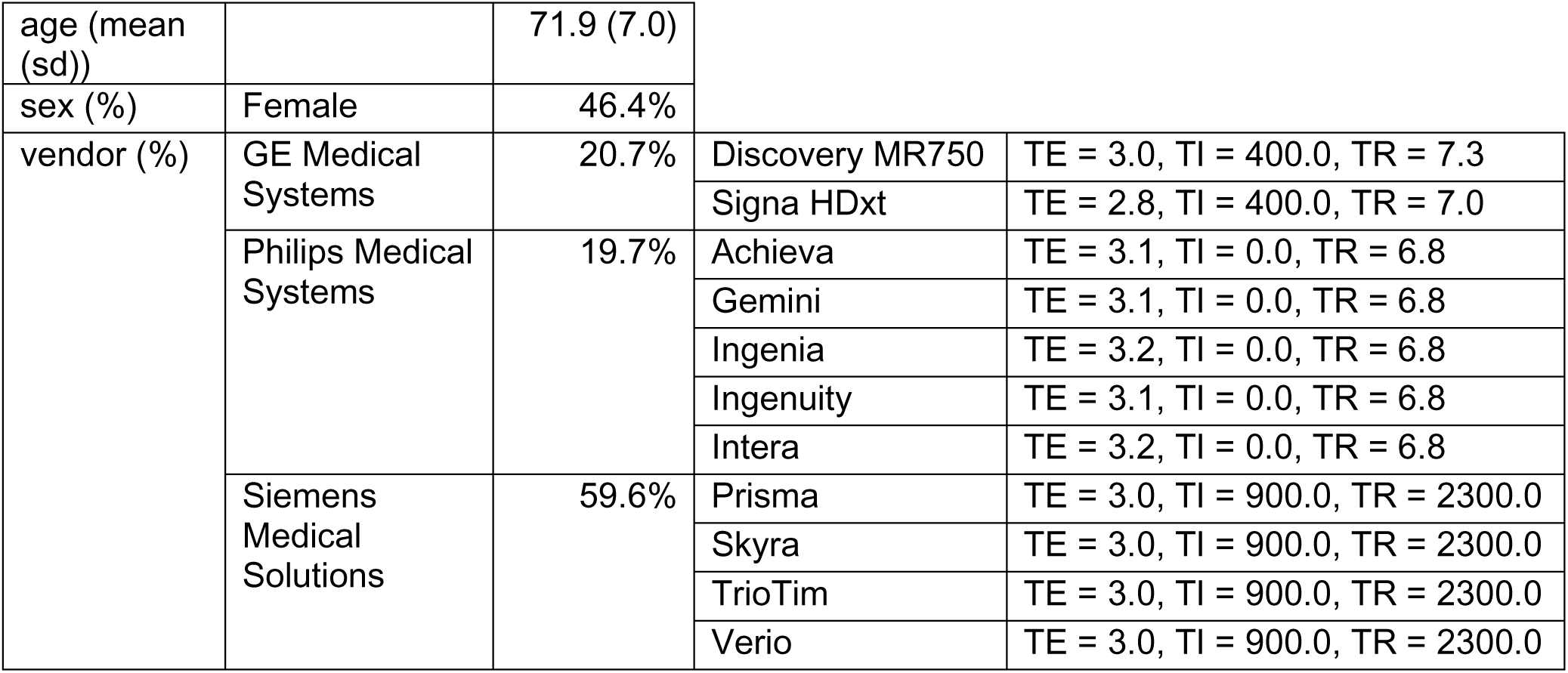
Demographic and scanner-related characteristics of the dataset, and acquisition details in ADNI. Values are reported as mean (standard deviation) for age, as percentages for categorical variables (sex and scanner manufacturer), and in milliseconds for acquisition parameters.

|  |  |  |  |  |
| --- | --- | --- | --- | --- |
| age (mean (sd)) |  | 71.9 (7.0) |  |  |
| sex (%) | Female | 46.4% |  |  |
| vendor (%) | GE Medical Systems | 20.7% | Discovery MR750 | TE = 3.0, TI = 400.0, TR = 7.3 |
|  |  |  | Signa HDxt | TE = 2.8, TI = 400.0, TR = 7.0 |
|  | Philips Medical Systems | 19.7% | Achieva | TE = 3.1, TI = 0.0, TR = 6.8 |
|  |  |  | Gemini | TE = 3.1, TI = 0.0, TR = 6.8 |
|  |  |  | Ingenia | TE = 3.2, TI = 0.0, TR = 6.8 |
|  |  |  | Ingenuity | TE = 3.1, TI = 0.0, TR = 6.8 |
|  |  |  | Intera | TE = 3.2, TI = 0.0, TR = 6.8 |
|  | Siemens Medical Solutions | 59.6% | Prisma | TE = 3.0, TI = 900.0, TR = 2300.0 |
|  |  |  | Skyra | TE = 3.0, TI = 900.0, TR = 2300.0 |
|  |  |  | TrioTim | TE = 3.0, TI = 900.0, TR = 2300.0 |
|  |  |  | Verio | TE = 3.0, TI = 900.0, TR = 2300.0 |

### 2.2. Artifact generation

We added four types of simulated artifacts (blurring, ghosting, motion, and noise) to the 1,000 MRI scans using TorchIO (Perez-Garcia et al., 2021). As TorchIO’s artifact simulations are dependent on image intensities, we normalized the mean and variance of each scan prior to applying the simulations. These normalized images will be referred to as non-degraded images. For each artifact type, we generated ten severity levels (**Figure 1**) by linearly adjusting the relevant parameters of the TorchIO functions. The lower and upper boundaries for each artifact’s parameter range (sections 2.1-2.4) were defined based on previous work (Loizillon et al., 2024), ensuring a clinically realistic spectrum. This approach resulted in 1,000 MRI scans × 4 artifact types × 10 severity levels = 40,000 degraded images, in addition to the 1000 non-degraded images.

**Figure 1.**
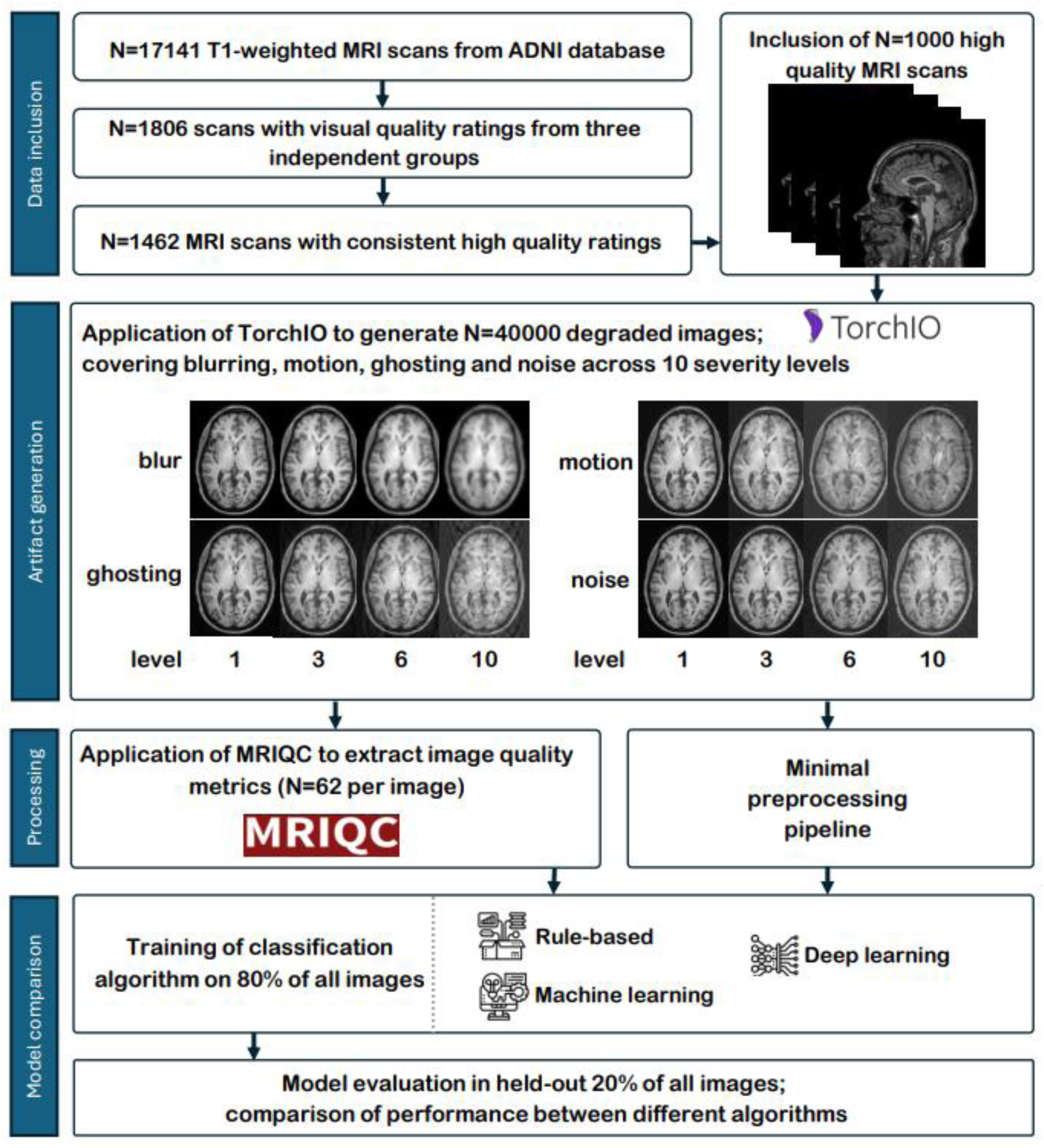
Schematic overview of methods.

#### 2.2.1 Blurring

TorchIO’s *RandomBlur* function applies a Gaussian kernel to blur the image along each axis (Perez-Garcia et al., 2021). By linearly increasing the standard deviations of the kernels (0.2 to 2) we created various levels of blurring artifact severity.

#### 2.2.2 Ghosting

TorchIO’s *RandomGhosting* function generates ghosting artifacts by Fourier transforming the image to k-space, creating several shifted copies of the k-space data along a specified axis, scaling these copies by a ghosting intensity parameter, and combining them with the original before inverse Fourier transforming back to the original space. We set the axis to anterior-posterior and linearly increased the ghosting intensity (0.15 to 1.5) for ten severity levels.

#### 2.2.3 Motion

TorchIO’s *RandomMotion* function (Richard Shaw, 2019), simulates rigid-body motion by applying a sequence of random rotations and translations in k-space (Perez-Garcia et al., 2021; Richard Shaw, 2019). The interpolation method was set to b-spline, and by linearly increasing both rotation (1 to 10 degrees) and translation parameters (1 to 10 mm) in tandem, we created ten severity levels.

#### 2.2.4 Noise

TorchIO’s *RandomNoise* function adds zero-mean Gaussian noise without spatial correlation (Santiago Aja-Fernández, 2016). By linearly increasing the noise standard deviations (0.03 to 0.3) we created ten severity levels. Noise values below 0 were retained.

### 2.3. Visual QC of degraded scans

To evaluate how the simulated artifacts relate to those typically observed in real-world scans, we conducted visual QC on a subset of the data using the PondrAI QC rating scale (1=perfect quality, 6 = terrible quality) (Costantino and Devenyi, 2023). 3,000 images were randomly selected from the 40,000 degraded images by shuffling the eligible image pool and then selecting the first 3,000, ensuring a representative distribution across all artifact types and severity levels (roughly 75 scans * 4 artifacts * 10 severity levels). Prior to rating, all 9 raters completed a standardized training protocol by reviewing the manual provided by PondrAI (Costantino and Devenyi, 2023) and practicing on a separate set of 100 images randomly selected from the remaining pool (excluding the 3,000 chosen images). A consensus meeting followed the practice session to align interpretations of the rating criteria. Each rater was assigned a subset of 1,000 images such that each image was rated by three raters.

We assessed inter-rater reliability using Krippendorff’s α and the average pairwise Spearman’s rank correlation coefficient (ρ) using R (version 4.4.1). Krippendorff’s α was chosen because it is suitable for ordinal data, accommodates missing values, and adjusts for agreement expected by chance, where agreement is indicated as ≥0.8 high, 0.67-0.79 moderate and <0.67 poor (Krippendorff, 2018). Spearman’s ρ was used to quantify the consistency of the relative ranking of images between raters, with correlations indicated as <0.39 weak, 0.40-0.59 moderate, 0.60-0.79 strong, and ≥0.8 very strong.

Krippendorff’s α was computed using the “ordinal” method, with 95% confidence intervals estimated via bootstrapping (1000 iterations). For Spearman’s ρ, all pairwise correlations between raters were calculated for each artifact type using the “*pairwise.complete.obs*” option to handle missing data, and then averaged (Nogueira et al., 2017). Confidence intervals for Spearman’s ρ were calculated using the Jackknife Euclidean likelihood-based inference approach (de Carvalho and Marques, 2012).

### 2.4. Model training

We evaluated three different types of artifact classification algorithms: RB, ML, and DL. For each artifact type and severity level, a separate model was trained to perform binary classification (e.g., the presence vs absence of blurring at severity level 5), resulting in 40 RB models, 40 ML models, and 40 DL models. No combinations of artifacts or severities were trained. All algorithms were implemented using scikit-learn (version 1.2.2).

To prevent data leakage, the data were split at the participant level, with all longitudinal scans from each participant (section 1) assigned exclusively to one set, resulting in a training set of 765 images and a test set of 235 images. Within the training set, 5-fold cross-validation was performed at the participant level: in each fold, 612 images (80% of the training set) were used for model training, and 153 images (20% of the training set) were used for validation, again ensuring that all data from a given participant remained within a single set. For each artifact type and severity level, separate datasets were created, resulting in a total of 40 datasets (4 artifact types × 10 severity levels). All three models were trained independently for each artifact and each severity level, resulting in 120 models.

#### 2.4.1. Input features RB and ML-based classification models

As input features for the RB and ML-based classification algorithms, we derived Image Quality Metrics (IQM) from the 1000 non-degraded and 40000 degraded images using MRIQC (version 22.0.1). Briefly, MRIQC performs head motion estimation with AFNI (version 22.0.17; Cox (1996)), followed by brain extraction with SynthStrip (version 1; Hoopes et al. (2022)), tissue segmentation with FSL FAST (version 5.0.11; Jenkinson et al. (2012)), and several image registration steps to MNI space with ANTS (version 2.3.3.dev168-g29bdf; Avants et al. (2008)), without accounting for dependencies between multiple sessions from the same participant. Subsequently, MRIQC derives a set of 68 IQMs based on the Quality Assessment Protocol (Shehzad et al., 2015). The six spatial resolution IQMs (x/y/z matrix size and voxel-size) were excluded because the spatial resolution does not depend on artifacts, resulting in 62 IQMs (**Supplementary Table 1**).

To distinguish normal from abnormal IQM values, we reuse predefined quality thresholds if they existed in previous literature, which was the case for 5 IQMs. These reference-based IQMs have a single absolute quality threshold (e.g., intracranial tissue volume fraction (Esteban et al., 2017)). The other 57 non-reference-based IQMs lack established thresholds and are typically used for relative quality comparisons (e.g., signal-to-noise ratio [SNR] (Esteban et al., 2017)). To harmonize vendor-based batch effects among the non-reference IQMs, we calculated the mean and standard deviation for each IQM per vendor in the 1000 non-degraded images, and used these to calculate IQM Z-scores for the 40000 degraded images.

#### 2.4.2 RB classification model

To develop and evaluate the RB classification model, we applied a multi-stage approach within each cross-validation fold, with the primary goal of determining optimal thresholds at the individual IQM level and then deriving a single threshold to classify the overall image quality. Thresholds for reference-based IQMs were fixed based on previously established values in MRIQC (Esteban et al., 2017). The non-reference-based IQM thresholds were determined within each training fold by evaluating a range of z-score thresholds for each non-reference IQM (from z = –5 to 5, in increments of 0.05), and selecting the threshold that maximized the Youden Index (YI; [specificity + sensitivity – 1]) in the validation set (Kim et al., 2019).

Subsequently, the reference-based IQM thresholds and optimized non-reference-based IQM thresholds were used to binarize IQMs in the training and validation subsets of that fold. Per image, the binary IQM values (1 good quality or 0 poor quality) were summed to a compound quality score, ranging from 0 (all IQMs flagged as poor quality) to 62 (all IQMs flagged as good quality). To define the final classification threshold for artifact presence, we used the compound scores from the non-degraded images within the training subset of that fold as a reference, and set the cut-off as the mean + 1 standard deviation.

#### 2.4.3 ML-based classification model

The ML-based classification algorithm used nested cross-validation to train and evaluate a Random Forest Classifier (RFC) (Esteban et al., 2017). Model training was performed using five outer folds for feature selection and model evaluation, and five inner folds for hyperparameter tuning.

To focus on the most informative IQMs while minimizing spurious or site-related influences, a two-step feature selection pipeline was applied in each outer fold (Esteban et al., 2017). First, an Extra Trees Classifier was trained to predict site using all IQMs (Geurts et al., 2006), and the IQMs most predictive of site were removed. Second, a Winnow-based selection was performed to retain only those features that provided more information than a randomly generated feature. Subsequently, this reduced feature set was used to optimize the inner folds hyperparameters.

Within each inner fold, a grid search was conducted to optimize the RFC’s hyperparameters for YI (number of estimators [10, 100, 500], maximum features [2, 4, 6, 8, 10, 15, 20], maximum depth [10, 20, 30, 50], and minimum samples split [5, 10, 20]).

#### 2.4.4 DL-based classification model

Unlike RB and ML-based classification models, which rely on IQMs extracted with MRIQC, the DL classification algorithms were directly trained on minimally preprocessed non-skull-stripped images. To keep the preprocessing comparable between the classification algorithms, we modified an existing preprocessing pipeline (Bottani et al., 2022) to correspond with the preprocessing of MRIQC. Bias field correction was applied using the N4ITK method (Tustison et al., 2010), and spatial registration to MNI was performed using ANTs. The minimum and maximum MNI space T1w image values were rescaled to a range ∈ [0,1].

We trained an existing convolutional neural network architecture with its original hyperparameters and loss functions (Bottani et al., 2022), using five convolutional blocks followed by three fully connected layers. Each convolutional block consisted of a convolutional layer, a batch normalization layer, a Rectified Linear Unit, and a max pooling layer. This CNN previously demonstrated the best performance for MRI QC in a large heterogeneous clinical dataset.

### 2.5. Final model evaluation

For all 120 models, the configuration (e.g., thresholds, hyperparameters, model weights) achieving the highest YI across all five folds was selected. Notably, for ML-based classification this involved the outer folds of the nested cross-validation procedure. Subsequently, the final model performance of each model was evaluated on the independent test set (n=235). In addition to our primary performance metric YI, we also calculated accuracy, balanced accuracy, F1 score, precision and recall to evaluate prediction accuracy.

### 2.6. Feature importance of RB– and ML models

To explore model interpretability of RB and ML algorithms, IQM importance was examined across the artifact types and severity levels. For each of the 80 RB and ML models, the most important IQMs were identified as those with the highest YI in RB models and those retained after feature selection in ML models, and these were recorded across all artifact and severity combinations for the final model.

For interpretation purposes, IQMs are grouped into four main categories: entropy, contrast, inhomogeneity and normative IQMs ((Alfaro-Almagro et al., 2018; Esteban et al., 2017; Hendriks et al., 2024; Shehzad et al., 2015). Entropy IQMs, such as the entropy focus criterion (EFC), capture the randomness or complexity in image intensity distributions. Image contrast IQMs, for example the contrast-to-noise ratio (CNR), quantify the distinguishability between different tissue types or structures. Intensity inhomogeneity IQMs, such as intensity non-uniformity (INU), assess the low frequency spatial uniformity of image intensity. Normative IQMs, including metrics like the overlap of tissue probability maps and the scan segmentations (TPM overlap), compare image characteristics to a reference population or template to detect deviations from expected norms. For each artifact type, Spearman’s rank correlations (⍴) were computed between severity levels (ordinal, 1 – 10) and IQM values (EFC, CNR, INU, TMP). ⍴ values were interpreted according to the following bins: negligible (< |0.10|), weak (|0.10| – |0.39|), moderate (|0.40-0.69|), strong (|0.70-0.89|), and very strong (|>0.90|) (Akoglu, 2018).

## 3. Results

### 3.1. Dataset characteristics

Demographic characteristics of the study population, including age, sex, and scanner vendor distribution for the 1,000 non-degraded images are summarized in **Table 1**. Descriptive statistics of the 62 IQMs of these images, stratified by vendor, are provided in **Supplementary Table 1**.

### 3.2. Visual QC ratings across simulated artifacts

Visual QC ratings increased with artifact severity for most artifact types (**Figure 2**). Overall, higher simulated artifact severity was associated with higher QC ratings. *Blurring* and *ghosting* showed a trend of linear increase across severity levels, indicating that simulated severity corresponded well to the PondrAI QC rating scale (i.e., higher artifact severity was associated with poorer image quality). For *motion*, ratings reported a logarithmic trend, with intermediate motion severities already scoring high on the PondrAI QC scale. *Noise* showed only a slight upward trend, with most ratings remaining between 2 and 4.

**Figure 2.**
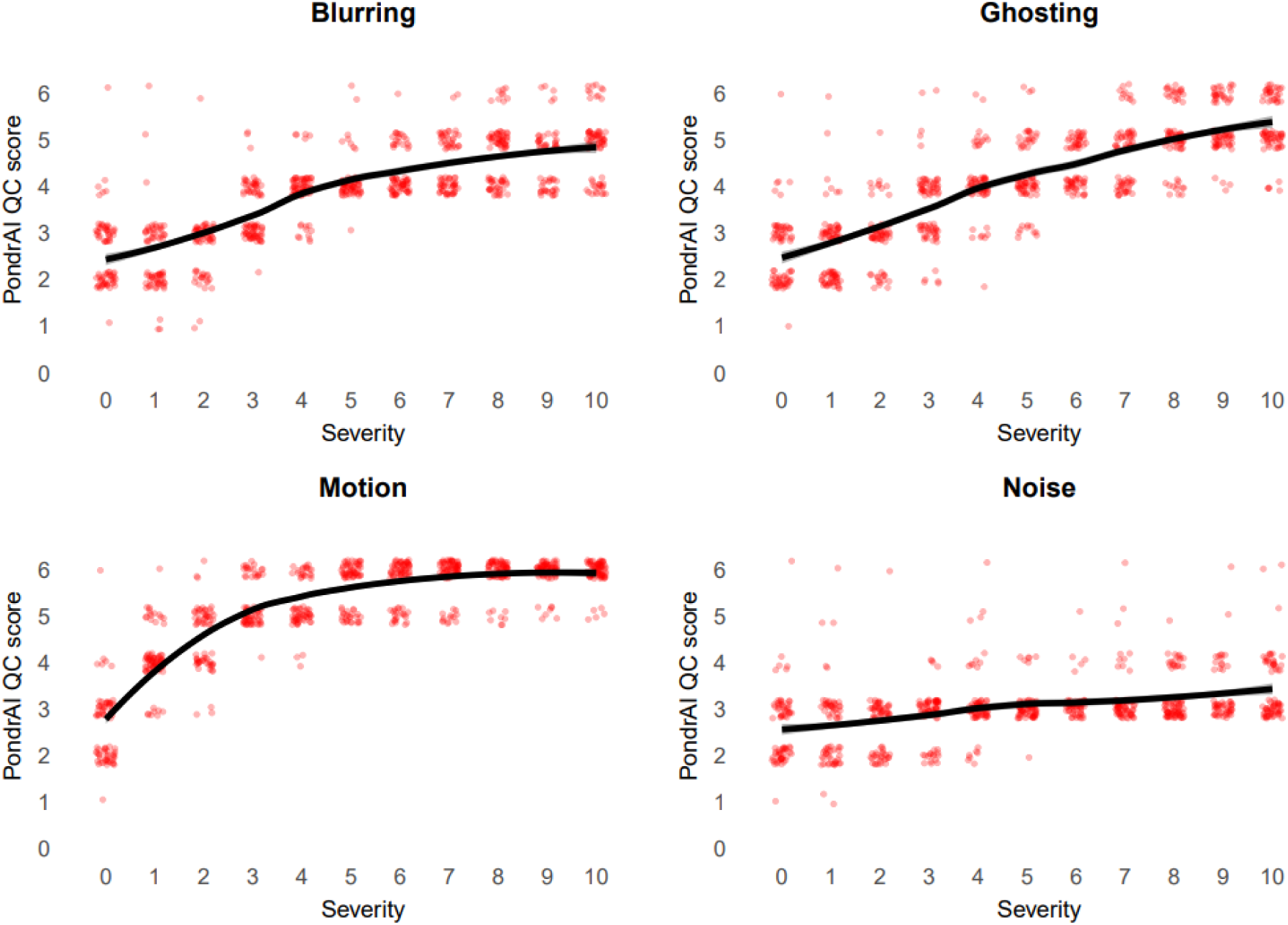
PondrAI QC ratings across artifact severities. Visual QC ratings based on the PondrAI QC scale (Costantino and Devenyi, 2023) as a function of artifact severity (0–10) for four artifact types: blurring, ghosting, motion, and noise. Red dots show individual ratings; black lines show smoothed trend lines with 95% confidence intervals, computed using locally estimated scatterplot smoothing (LOESS). Blurring and ghosting display an approximately linear increase in QC ratings with severity, motion shows a ceiling effect, and noise shows minimal change.

While inter-rater reliability was strong when assessed across all artifact types (Krippendorff’s α = 0.82, 95% CI: 0.81–0.83; mean Spearman’s ρ = 0.87, 95% CI: 0.84–0.90), it varied considerably between individual artifacts. *Ghosting* showed the highest agreement (Krippendorff’s α = 0.79, 95% CI: 0.77–0.82; mean Spearman’s ρ = 0.84, 95% CI: 0.76–0.92). *Blurring* and *motion* had poor agreement based on Krippendorff’s α (0.63, 95% CI: 0.59–0.66 and 0.54, 95% CI: 0.49–0.58 respectively), but still relatively high consistency in ranking (mean Spearman’s ρ = 0.77, 95% CI: 0.65–0.88 and mean Spearman’s ρ = 0.73, 95% CI: 0.57–0.88 respectively). For both *noise* and *non-degraded images*, reliability was lower, with Krippendorff’s α values of 0.41 (95% CI: 0.35–0.46) and 0.31 (95% CI: 0.13–0.45) and lower mean Spearman’s ρ values (0.46, CI: 0.26–0.70 and 0.32, CI: 0.55–0.87), indicating less agreement in both scores and image ranking. These differences are further illustrated in the pairwise Spearman’s ρ heatmaps (**Supplementary Figure 1**).

### 3.3. RB, ML, and DL model comparisons

Detailed RB, ML, DL-based classification results for blurring, ghosting, noise, and motion and severity levels 1–10 are shown in **Table 2**.

**Table 2.** Performance metrics of rule-based, machine learning based and deep learning based models for the different artifacts and severities.

| artifact | severity | balanced accuracy |  |  | accuracy |  |  | f1 score |  |  | precision |  |  | recall |  |  | Youden Index |  |  |
| --- | --- | --- | --- | --- | --- | --- | --- | --- | --- | --- | --- | --- | --- | --- | --- | --- | --- | --- | --- |
|  |  | RB | ML | DL | RB | ML | DL | RB | ML | DL | RB | ML | DL | RB | ML | DL | RB | ML | DL |
| blurring | 1 | 0.51 | 0.50 | 0.92 | 0.51 | 0.50 | 0.92 | 0.62 | 0.51 | 0.92 | 0.19 | 0.49 | 0.86 | 0.82 | 0.52 | 0.98 | 0.01 | 0.00 | 0.83 |
| blurring | 2 | 0.65 | 0.61 | 0.92 | 0.65 | 0.61 | 0.92 | 0.70 | 0.56 | 0.92 | 0.51 | 0.71 | 0.88 | 0.79 | 0.50 | 0.96 | 0.30 | 0.22 | 0.84 |
| blurring | 3 | 0.88 | 0.99 | 0.98 | 0.88 | 0.99 | 0.98 | 0.87 | 0.99 | 0.98 | 0.94 | 0.99 | 0.97 | 0.82 | 1.00 | 1.00 | 0.75 | 0.99 | 0.97 |
| blurring | 4 | 0.93 | 1.00 | 0.99 | 0.93 | 1.00 | 0.99 | 0.93 | 1.00 | 0.99 | 1.00 | 1.00 | 0.98 | 0.87 | 1.00 | 1.00 | 0.86 | 1.00 | 0.98 |
| blurring | 5 | 0.90 | 1.00 | 0.99 | 0.90 | 1.00 | 0.99 | 0.89 | 1.00 | 0.99 | 1.00 | 1.00 | 0.97 | 0.80 | 1.00 | 1.00 | 0.80 | 1.00 | 0.97 |
| blurring | 6 | 0.90 | 1.00 | 0.98 | 0.90 | 1.00 | 0.98 | 0.89 | 1.00 | 0.98 | 1.00 | 1.00 | 0.97 | 0.81 | 1.00 | 1.00 | 0.80 | 1.00 | 0.97 |
| blurring | 7 | 0.90 | 1.00 | 0.98 | 0.90 | 1.00 | 0.98 | 0.89 | 1.00 | 0.98 | 1.00 | 1.00 | 0.97 | 0.80 | 1.00 | 1.00 | 0.80 | 1.00 | 0.97 |
| blurring | 8 | 0.89 | 1.00 | 0.98 | 0.89 | 1.00 | 0.98 | 0.87 | 1.00 | 0.98 | 1.00 | 1.00 | 0.95 | 0.77 | 1.00 | 1.00 | 0.77 | 1.00 | 0.95 |
| blurring | 9 | 0.88 | 1.00 | 0.98 | 0.88 | 1.00 | 0.98 | 0.87 | 1.00 | 0.98 | 1.00 | 1.00 | 0.96 | 0.77 | 1.00 | 1.00 | 0.77 | 1.00 | 0.95 |
| blurring | 10 | 0.90 | 1.00 | 0.99 | 0.90 | 1.00 | 0.99 | 0.89 | 1.00 | 0.99 | 1.00 | 1.00 | 0.97 | 0.81 | 1.00 | 1.00 | 0.81 | 1.00 | 0.97 |
| ghosting | 1 | 0.53 | 0.52 | 0.92 | 0.53 | 0.52 | 0.92 | 0.61 | 0.54 | 0.93 | 0.32 | 0.48 | 0.85 | 0.74 | 0.56 | 1.00 | 0.07 | 0.04 | 0.85 |
| ghosting | 2 | 0.57 | 0.57 | 0.95 | 0.57 | 0.57 | 0.95 | 0.64 | 0.58 | 0.95 | 0.38 | 0.54 | 0.92 | 0.76 | 0.60 | 0.98 | 0.14 | 0.14 | 0.90 |
| ghosting | 3 | 0.65 | 0.65 | 0.95 | 0.65 | 0.65 | 0.95 | 0.70 | 0.66 | 0.95 | 0.51 | 0.62 | 0.93 | 0.80 | 0.68 | 0.97 | 0.30 | 0.30 | 0.90 |
| ghosting | 4 | 0.73 | 0.71 | 0.95 | 0.73 | 0.71 | 0.95 | 0.75 | 0.72 | 0.95 | 0.65 | 0.69 | 0.91 | 0.81 | 0.74 | 0.99 | 0.46 | 0.43 | 0.90 |
| ghosting | 5 | 0.80 | 0.80 | 0.94 | 0.80 | 0.80 | 0.94 | 0.81 | 0.81 | 0.94 | 0.78 | 0.79 | 0.88 | 0.83 | 0.82 | 1.00 | 0.61 | 0.61 | 0.88 |
| ghosting | 6 | 0.82 | 0.87 | 0.94 | 0.82 | 0.87 | 0.94 | 0.81 | 0.87 | 0.95 | 0.83 | 0.87 | 0.89 | 0.80 | 0.87 | 1.00 | 0.63 | 0.73 | 0.89 |
| ghosting | 7 | 0.85 | 0.90 | 0.95 | 0.85 | 0.90 | 0.95 | 0.84 | 0.90 | 0.95 | 0.87 | 0.91 | 0.91 | 0.82 | 0.90 | 0.98 | 0.70 | 0.81 | 0.89 |
| ghosting | 8 | 0.89 | 0.91 | 0.95 | 0.89 | 0.91 | 0.95 | 0.88 | 0.92 | 0.95 | 0.93 | 0.91 | 0.92 | 0.85 | 0.92 | 0.98 | 0.78 | 0.83 | 0.90 |
| ghosting | 9 | 0.88 | 0.94 | 0.96 | 0.88 | 0.94 | 0.96 | 0.87 | 0.94 | 0.96 | 0.95 | 0.91 | 0.92 | 0.81 | 0.96 | 1.00 | 0.75 | 0.87 | 0.92 |
| ghosting | 10 | 0.89 | 0.96 | 0.96 | 0.89 | 0.96 | 0.96 | 0.88 | 0.96 | 0.96 | 0.99 | 0.95 | 0.93 | 0.79 | 0.97 | 0.99 | 0.78 | 0.92 | 0.92 |
| motion | 1 | 0.74 | 0.67 | 0.96 | 0.74 | 0.67 | 0.96 | 0.76 | 0.66 | 0.97 | 0.62 | 0.71 | 0.93 | 0.85 | 0.63 | 1.00 | 0.47 | 0.34 | 0.93 |
| motion | 2 | 0.77 | 0.67 | 0.96 | 0.77 | 0.67 | 0.96 | 0.78 | 0.68 | 0.96 | 0.70 | 0.67 | 0.91 | 0.84 | 0.68 | 1.00 | 0.54 | 0.35 | 0.91 |
| motion | 3 | 0.90 | 0.94 | 0.98 | 0.90 | 0.94 | 0.98 | 0.89 | 0.94 | 0.98 | 0.98 | 0.94 | 0.96 | 0.83 | 0.95 | 1.00 | 0.80 | 0.89 | 0.95 |
| motion | 4 | 0.89 | 0.90 | 0.96 | 0.89 | 0.90 | 0.96 | 0.88 | 0.90 | 0.96 | 0.94 | 0.87 | 0.93 | 0.84 | 0.92 | 1.00 | 0.78 | 0.80 | 0.92 |
| motion | 5 | 0.90 | 0.93 | 0.96 | 0.90 | 0.93 | 0.96 | 0.89 | 0.94 | 0.96 | 0.98 | 0.91 | 0.93 | 0.82 | 0.96 | 0.99 | 0.80 | 0.87 | 0.91 |
| motion | 6 | 0.91 | 0.97 | 0.95 | 0.91 | 0.97 | 0.95 | 0.90 | 0.97 | 0.95 | 1.00 | 0.96 | 0.92 | 0.82 | 0.97 | 0.99 | 0.82 | 0.93 | 0.91 |
| motion | 7 | 0.92 | 0.95 | 0.96 | 0.92 | 0.95 | 0.96 | 0.91 | 0.95 | 0.96 | 0.99 | 0.96 | 0.94 | 0.85 | 0.94 | 0.99 | 0.84 | 0.89 | 0.93 |
| motion | 8 | 0.90 | 0.97 | 0.96 | 0.90 | 0.97 | 0.96 | 0.89 | 0.97 | 0.96 | 1.00 | 0.97 | 0.94 | 0.81 | 0.96 | 0.98 | 0.81 | 0.94 | 0.92 |
| motion | 9 | 0.90 | 0.96 | 0.97 | 0.90 | 0.96 | 0.97 | 0.88 | 0.96 | 0.97 | 1.00 | 0.96 | 0.95 | 0.79 | 0.96 | 0.99 | 0.79 | 0.92 | 0.94 |
| motion | 10 | 0.90 | 0.98 | 0.98 | 0.90 | 0.98 | 0.98 | 0.89 | 0.98 | 0.98 | 1.00 | 0.98 | 0.96 | 0.80 | 0.99 | 1.00 | 0.80 | 0.97 | 0.96 |
| noise | 1 | 0.50 | 0.53 | 0.94 | 0.50 | 0.53 | 0.94 | 0.63 | 0.55 | 0.94 | 0.16 | 0.51 | 0.90 | 0.84 | 0.56 | 0.97 | 0.00 | 0.07 | 0.88 |
| noise | 2 | 0.55 | 0.60 | 0.97 | 0.55 | 0.60 | 0.97 | 0.64 | 0.60 | 0.97 | 0.27 | 0.60 | 0.97 | 0.82 | 0.60 | 0.97 | 0.09 | 0.20 | 0.94 |
| noise | 3 | 0.62 | 0.72 | 0.97 | 0.62 | 0.72 | 0.97 | 0.69 | 0.71 | 0.98 | 0.41 | 0.72 | 0.95 | 0.83 | 0.71 | 1.00 | 0.24 | 0.43 | 0.95 |
| noise | 4 | 0.70 | 0.89 | 0.97 | 0.70 | 0.89 | 0.97 | 0.73 | 0.89 | 0.97 | 0.60 | 0.89 | 0.95 | 0.80 | 0.89 | 0.99 | 0.40 | 0.78 | 0.94 |
| noise | 5 | 0.78 | 0.95 | 0.97 | 0.78 | 0.95 | 0.97 | 0.79 | 0.95 | 0.97 | 0.74 | 0.94 | 0.94 | 0.83 | 0.96 | 1.00 | 0.57 | 0.90 | 0.94 |
| noise | 6 | 0.85 | 0.98 | 0.98 | 0.85 | 0.98 | 0.98 | 0.85 | 0.98 | 0.98 | 0.84 | 0.97 | 0.95 | 0.85 | 0.98 | 1.00 | 0.69 | 0.95 | 0.95 |
| noise | 7 | 0.89 | 0.99 | 0.97 | 0.89 | 0.99 | 0.97 | 0.89 | 0.99 | 0.97 | 0.93 | 0.98 | 0.94 | 0.85 | 0.99 | 1.00 | 0.78 | 0.97 | 0.94 |
| noise | 8 | 0.91 | 1.00 | 0.97 | 0.91 | 1.00 | 0.97 | 0.91 | 1.00 | 0.97 | 0.96 | 0.99 | 0.94 | 0.86 | 1.00 | 1.00 | 0.82 | 0.99 | 0.94 |
| noise | 9 | 0.94 | 1.00 | 0.97 | 0.94 | 1.00 | 0.97 | 0.94 | 1.00 | 0.98 | 0.99 | 1.00 | 0.95 | 0.89 | 1.00 | 1.00 | 0.88 | 1.00 | 0.95 |
| noise | 10 | 0.95 | 1.00 | 0.98 | 0.95 | 1.00 | 0.98 | 0.95 | 1.00 | 0.98 | 1.00 | 1.00 | 0.97 | 0.90 | 1.00 | 1.00 | 0.90 | 1.00 | 0.97 |

For *blurring*, DL-based classification demonstrated strong discrimination across all severities (YI = 0.83 – 0.97, **Figure 3**). RB classification showed a strong upward trend, especially at severity 3, with YI rising from 0.30 to 0.75 and continuing to improve to very strong performance (YI = 0.81) at the highest severity. ML-based performance was also initially weak with YI = 0 at severity 1, but improved rapidly, reaching near-perfect levels (YI ≥ 0.99) by severity 3. For *ghosting*, DL models maintained strong discrimination (YI 0.85–0.92) across all severities, while both RB and classical ML models showed weak discrimination at early severities (YI<0.15), improving to strong (YI = 0.78) and very strong (YI = 0.92) discrimination, respectively, at maximum severity. For *motion*, DL based classification achieved consistently very strong YI (0.91–0.96), while RB models showed moderate discrimination at low severity (0.47), improving to 0.80 at severity 10, and classical ML based models exhibited a similar trend but with higher maximum performance (YI=0.97). In the case of *noise*, DL classification maintained strong performance with YI between 0.88 and 0.97, outperforming both RB and ML models at lower severities; classical ML reached perfect discrimination (YI = 1.00) at higher severities, whereas RB models improved gradually from 0.00 to 0.90.

**Figure 3.**
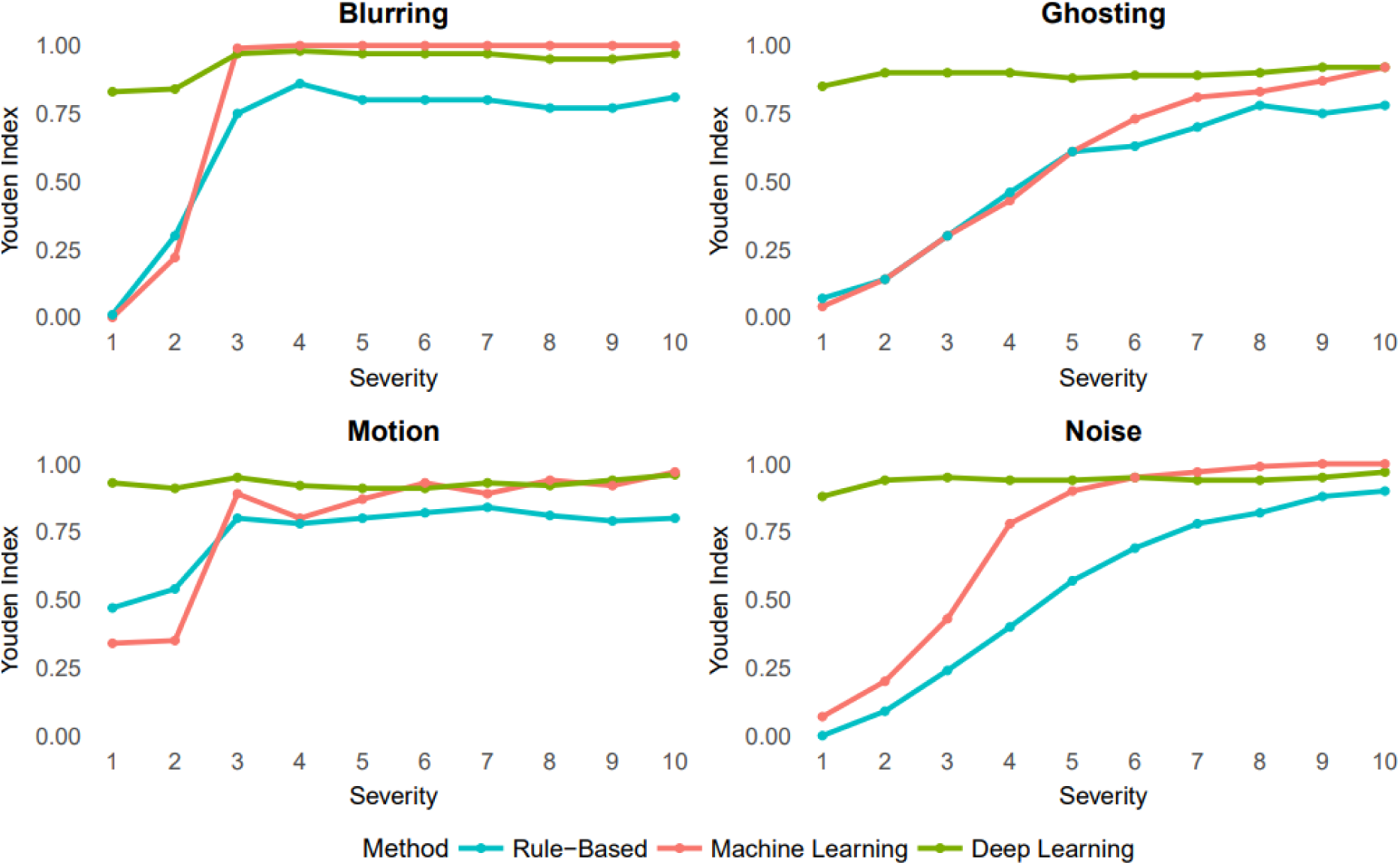
Youden Index scores across artifact severity levels (1–10) for each artifact type (blur, ghosting, motion, and noise), comparing performance of rule-based (blue), machine learning-based (red), and deep learning-based (green) classification methods. Each panel presents results for a single artifact type. Lines represent the Youden Index (sensitivity + specificity – 1) at each severity level.

Overall, these findings indicate that DL-based classification is consistently robust across all artifact types and severities, while RB and ML models are more reliably performant only at higher levels of artifact severity and show limited discrimination accuracy at lower severities across all artifact types. Other evaluation metrics, displaying the same trends as the YI, are shown in **Supplementary Figures 2-6**.

### 3.4. IQM importance for RB and ML-based classification

IQMs exhibited distinct patterns across artifact types and severity levels (**Supplementary Figure 7**, **Supplementary Table 2**). For *blurring*, there were moderately increased values of EFC (⍴ = 0.68) and CNR (⍴ = 0.56), and a weak positive association for TPM (⍴ = 0.33), alongside a weak negative association for INU (ρ = –0.24), indicating greater image entropy, enhanced contrast, and tissue probability map distortion at higher severities. *Ghosting* led to weak increases in INU (ρ = 0.35) and weak declines in TPM (ρ = –0.31), while EFC and CNR remained relatively stable (ρ = –0.15 and ρ = 0.05), suggesting spatial misalignment without major changes in contrast or entropy. *Motion* was associated with weakly increasing INU (ρ = 0.34) and moderately decreasing TPM (ρ = –0.42), reflecting spatial distortion and misclassification of tissue classes, while EFC and CNR remained stable. In contrast, *noise* resulted in declines in EFC (ρ = –0.71) and moderate increases in INU (ρ = 0.50), while showing negligible-to-weak declines in both CNR (ρ = –0.21) and TPM (ρ = –0.14). These trends highlight the complementary sensitivity of EFC, CNR, INU, and TPM to different artifact types and intensities (**Supplementary Figure 7**).

The pattern of IQM selection across artifact types and severity levels for the RB and ML-based classification is displayed in **Figure 4**. For *blurring*, both ML and RB models initially selected a broad mix of normative and entropy IQMs, alongside some image contrast IQMs. From severity level 3 onward, RB and ML models consistently prioritized normative and entropy IQMs, particularly those reflecting spatial properties and deviations in CSF, while rule-based models reduced their selection to a smaller set of normative features, mainly related to CSF and spatial structure. For *ghosting*, RB models relied mainly on normative IQMs related to the gray matter and white matter, and a few image contrast IQMs. ML models relied at lower severities mainly on normative IQMS related to the CSF, while at higher severities it shifted to normative IQMs related to the white matter, and some image contrast IQMs. For *motion*, both RB and ML models initially used normative IQMs. However RB models’ most informative normative IQMs were gray matter, white matter, and CSF related, whereas ML models used mainly background related normative IQMs. At higher severities the most informative IQMs for RB models were the normative IQMs. ML models relied on the GM and background related normative IQMs and SNR-related image contrast IQMs. For *noise*, ML models initially drew from all four categories but shifted toward normative IQMs, particularly those reflecting CSF, gray matter, and entropy measures (FWHM related). In contrast, RB models narrowed their focus more abruptly, retaining mainly normative (GM related) and image contrast (SNR related) IQMs.

**Figure 4.**
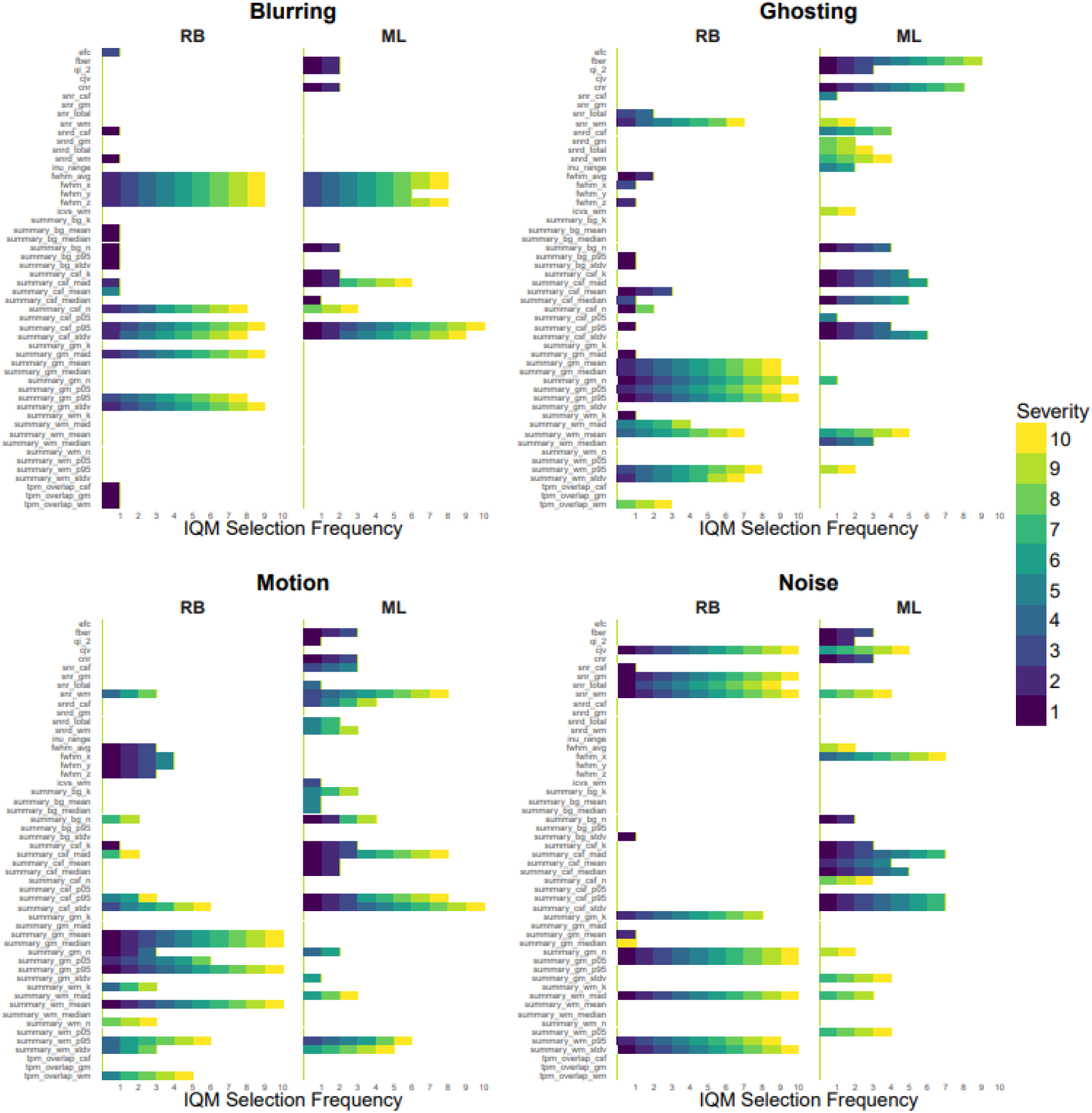
Frequency of IQM selection across artifact types, severity levels and classification models. Each panel shows the number of models (x-axis) that selected each IQM (y-axis) for a given artifact type (blur, ghosting, motion, noise), separately for rule-based (RB) and machine learning-based (ML) classification. Color indicates artifact severity (1–10).

## 4. Discussion

This benchmark study yielded three main findings. First, while visual QC ratings correlated with simulated artifact severity, the non-linearity of these relations varied between the artifacts. Second, while there was little performance difference for detecting high severity artifacts, DL outperformed RB and ML for detecting subtle artifacts. RB and ML performed similarly, with performance varying between artifact types. Thirdly, RB models relied mainly on a consistent core of normative features with limited use of contrast measures, whereas ML models adapted their feature use depending on artifact type and severity, drawing more flexibly from multiple IQM categories.

Our findings highlight that human visual QC is not equally effective across artifact types, which may have important implications for its use as a benchmark. The high inter-reader agreement for ghosting suggests that artifacts of this type can be reliably perceived. For blurring and motion, raters agreed on less but were still relatively consistent in the order in which they judged images from better to worse quality. Noise, in particular, was rated differently by different readers, perhaps reflecting its weaker perceptual salience, or limited noise variability in the higher severities of the PondrAIQC training examples. These findings are in line with previous studies. When comparing human visual QC with machine-based, non-artifact specific, QC scores, previous studies also found poor correspondence, particularly for subtle or borderline real-world artifacts (Pizarro et al., 2016; Rosen et al., 2018). Another study with synthetic artifacts found stronger correlations of radiologist diagnostic quality scores with perceptually aligned image quality metrics, such as DISTS and HaarPSI, than with other IQMs (Kastryulin et al., 2023). Together, this highlights the limitations of visual QC as a gold standard for subtle (synthetic) artifacts, whether for detecting general image quality or for detecting specific artifacts.

The fact that DL mostly outperformed RB and ML at low artifact severities aligns with previous studies, which state as explanation that DL architectures can learn more complex hierarchical image representations and more subtle textural and structural deviations than traditional image features (Bhaleraoa et al., 2025; Bottani et al., 2022; Fantini et al., 2021; Keshavan et al., 2019). This advantage seems especially pronounced for artifacts with distributed, non-local, changes in image intensity and spatial coherence, such as ghosting and motion (Esteban et al., 2017; Shaw et al., 2019). Nevertheless, RB and ML approaches remained highly effective in detecting blurring and noise artifacts, especially at higher severity levels, where degradation can be more easily captured by single or composite IQMs (Kim et al., 2019; Mortamet et al., 2009). In line with this, it has been shown that, in head-to-head comparison, classical ML models trained on robust IQMs can match DL models for motion artifacts (Vakli et al., 2023). Together with these studies, our findings indicate that simpler ML models offer a practical alternative to DL, which can be beneficial as they can be more efficient and easier to implement, especially when the goal is to merely detect gross artifacts.

The importance of IQMs for artifact detection by RB and ML varied with the artifact type and severity. At low artifact severities, both RB and ML relied on a broad spectrum of normative and image contrast IQMs to detect subtle or diffuse degradations. With increasing severity, the IQM selection consistently shifted toward normative features, particularly those quantifying deviations from the expected anatomical patterns and spatial properties. Similarly, previous studies demonstrated that global entropy and contrast IQMs are sensitive to early or mild artifacts (Alfaro-Almagro et al., 2018; Mortamet et al., 2009; Woodard and Carley-Spencer, 2006), while normative, region-based IQMs offer superior sensitivity and specificity for more severe or localized degradations (Esteban et al., 2017; Pizarro et al., 2016). The increasing reliance on normative IQMs is particularly relevant in large neuroimaging studies, where heterogeneity in acquisition and (physio-)pathology can obscure subtle artifacts unless deviations from normative patterns are explicitly modeled (Alfaro-Almagro et al., 2018; Esteban et al., 2017; Rosen et al., 2018). These results underscore the value of combining global and normative IQMs for comprehensive and robust MRI quality control using RB or ML.

A key strength of our study is the use of synthetic artifacts for benchmarking, which allows for precise control over artifact type and severity, and the ability to generate large, balanced datasets for model training and evaluation (Loizillon et al., 2024; Perez-Garcia et al., 2021). However, the generalizability of synthetic artifacts to real-world MRI artifacts may be limited, as these do not capture the heterogeneous and subtle degradations that arise from the complex interactions between head motion, hardware imperfections, and acquisition parameters (Bhaleraoa et al., 2025; Smith, 2010). These complex interactions likely lead to the presence of multiple artifact types, perhaps often as too low severities to be visible to the human eye. Moreover, real MRI scans often exhibit multiple, overlapping artifact types that can impact image quality simultaneously, perhaps often as too low severities to be visible to the human eye. Within this study, we chose to only compare the detectability of individual simulated artifacts to maximize interpretability. Perhaps, DL would outperform RB and ML even more when several synthetic artifacts are combined.

Recent work employed transfer learning to retrain DL models on small sets of real, artifact-labeled images after pretraining on synthetic artifacts (Loizillon et al., 2024), showing that this combination of simulated and real-world artifacts leads to higher artifact detection performance and generalizability. Although this combination of simulated and real-world artifacts led to higher performance and generalizability, it doesn’t allow disentangling the presence of individual artifacts. Automatic disentangling between artifact types may have value to differentiate scanner and sequence stability from patient compliance. Together, these findings suggest that future research should employ advanced explainable DL algorithms, trained with a combination of individual synthetic artifacts and real-world artifacts.

Another factor limiting the generalizability of our study is the reliance on a single dataset (ADNI) for both training and validation. Although ADNI is a multisite study, its standardized protocols and participant demographics may not fully represent the diversity encountered in clinical practice or other research cohorts (Alfaro-Almagro et al., 2018; Hendriks et al., 2024; Kruggel et al., 2010). This homogeneity could inflate model performance and limit its generalizability. Previous studies have highlighted the challenges of cross-site generalization for automated QC, emphasizing the need for diverse, multi-cohort benchmarks and harmonization strategies (Bhaleraoa et al., 2025; Esteban et al., 2017; White et al., 2018). Future studies should prioritize external validation using datasets with natural artifacts and broader demographic variability.

Furthermore, we prioritized comparability over performance, by employing ML and DL models that were often used and available open-source, with their standard parameter settings from previous studies. While these models ranked among the best-performing approaches for T1w QC (Bhaleraoa et al., 2025; Esteban et al., 2017; Fantini et al., 2021; Pizarro et al., 2016; Rosen et al., 2018), they may be outperformed by more recent and advanced methods, such as a ProtoPNet (Garcia et al., 2024) or by more extensive hyperparameter optimization or ensemble methods. Additionally, we did not investigate why DL models were more successful at detecting subtle artifacts, as exploring model interpretability, e.g., through saliency mapping, SHAP values or feature attribution methods (Bottani et al., 2022; Lundervold and Lundervold, 2019), was beyond the scope of this study. Systematically benchmarking advanced DL models and integrating explainable AI in future studies could establish the true upper bounds of automated QC performance in neuroimaging.

In conclusion, our findings demonstrate that DL offers superior performance for the automated detection of individual synthetic artifacts in T1w brain MRI, particularly for subtle artifacts. RB and ML methods may remain valuable for interpreting quality degradation of artifacts with well-defined quantifiable features. For RB and ML methods, the varying importance of IQMs across artifact types and severities highlight the need for flexible, multi-metric QC frameworks. Future studies should address the challenges of real-world generalizability, model complexity, and interpretability between synthetic and real-world artifacts to fully realize the potential of automated QC in neuroimaging.

## Data Availability

The code for artifact simulation and the training of the rule-based algorithm is available at the Corresponding author's GitHub: https://github.com/janinehendriks-hub/AutoQC-compare-MRI
The code for the machine-learning based method is available at: https://pypi.org/project/mriqc-learn/0.0.2/
The code for the deep-learning based method is available at: https://clinicadl.readthedocs.io/en/stable/

https://adni.loni.usc.edu/

## 5. Acknowledgements

This project was funded by Health∼Holland, Top Sector Life Sciences & Health (TKI-PPP Amsterdam UMC 2011227).

Data collection and sharing for the Alzheimer’s Disease Neuroimaging Initiative (ADNI) is funded by the National Institute on Aging (National Institutes of Health Grant U19AG024904). The grantee organization is the Northern California Institute for Research and Education. In the past, ADNI has also received funding from the National Institute of Biomedical Imaging and Bioengineering, the Canadian Institutes of Health Research, and private sector contributions through the Foundation for the National Institutes of Health (FNIH) including generous contributions from the following: AbbVie, Alzheimer’s Association; Alzheimer’s Drug Discovery Foundation; Araclon Biotech; BioClinica, Inc.; Biogen; Bristol-Myers Squibb Company; CereSpir, Inc.; Cogstate; Eisai Inc.; Elan Pharmaceuticals, Inc.; Eli Lilly and Company; EuroImmun; F. Hoffmann-La Roche Ltd and its affiliated company Genentech, Inc.; Fujirebio; GE Healthcare; IXICO Ltd.; Janssen Alzheimer Immunotherapy Research & Development, LLC.; Johnson & Johnson Pharmaceutical Research & Development LLC.; Lumosity; Lundbeck; Merck & Co., Inc.; Meso Scale Diagnostics, LLC.; NeuroRx Research; Neurotrack Technologies; Novartis Pharmaceuticals Corporation; Pfizer Inc.; Piramal Imaging; Servier; Takeda Pharmaceutical Company; and Transition Therapeutics.

