## Supplemental Tables and Figures for "Automatic detection of simulated artifacts on T1w magnetic resonance images: comparing performance of different QC strategies"

**Supplementary Table 1**. Mean and standard deviation (SD) of all 62 included IQMs across all MRI scans and by scanner manufacturer (GE Medical Systems, Philips Medical Systems, Siemens). IQMs are grouped according to their measurement domain: entropy, image contrast, intensity inhomogeneity and normative IQMs.

| IQM | All (Mean ± SD) | Ge Medical Systems (Mean ± SD) | Philips Medical Systems (Mean ± SD) | Siemens (Mean ± SD) |
|---|---|---|---|---|
| cjv | 0.48 ± 0.10 | 0.43 ± 0.06 | 0.38 ± 0.04 | 0.54 ± 0.09 |
| cnr | 1.94 ± 1.20 | 2.67 ± 1.18 | 2.90 ± 0.94 | 1.37 ± 0.90 |
| efc | 0.70 ± 0.04 | 0.65 ± 0.02 | 0.74 ± 0.01 | 0.70 ± 0.02 |
| fber | 42.67 ± 312.29 | 21.69 ± 54.56 | 29.79 ± 330.53 | 54.22 ± 355.51 |
| fwhm_avg | 3.90 ± 0.40 | 3.70 ± 0.24 | 4.59 ± 0.26 | 3.74 ± 0.18 |
| fwhm_x | 3.65 ± 0.39 | 3.29 ± 0.24 | 4.25 ± 0.34 | 3.57 ± 0.15 |
| fwhm_y | 4.28 ± 0.49 | 4.12 ± 0.28 | 5.12 ± 0.27 | 4.06 ± 0.23 |
| fwhm_z | 3.78 ± 0.37 | 3.69 ± 0.24 | 4.39 ± 0.24 | 3.60 ± 0.19 |
| icvs_csf | 0.25 ± 0.01 | 0.25 ± 0.01 | 0.26 ± 0.02 | 0.25 ± 0.01 |
| icvs_gm | 0.37 ± 0.02 | 0.38 ± 0.02 | 0.38 ± 0.02 | 0.37 ± 0.02 |
| icvs_wm | 0.38 ± 0.02 | 0.37 ± 0.01 | 0.37 ± 0.01 | 0.38 ± 0.01 |
| inu_med | 0.92 ± 0.06 | 0.94 ± 0.05 | 0.84 ± 0.06 | 0.94 ± 0.05 |
| inu_range | 0.17 ± 0.07 | 0.16 ± 0.06 | 0.26 ± 0.05 | 0.15 ± 0.06 |
| qi_1 | 0.00 ± 0.01 | 0.00 ± 0.01 | 0.00 ± 0.00 | 0.00 ± 0.00 |
| qi_2 | 0.06 ± 0.04 | 0.06 ± 0.04 | 0.10 ± 0.05 | 0.05 ± 0.03 |
| rpve_csf | 31.98 ± 3.82 | 36.11 ± 3.37 | 29.45 ± 3.04 | 31.38 ± 3.02 |
| rpve_gm | 19.39 ± 2.13 | 21.69 ± 1.71 | 18.11 ± 1.72 | 19.01 ± 1.77 |
| rpve_wm | 24.80 ± 3.40 | 29.16 ± 2.73 | 23.73 ± 2.46 | 23.63 ± 2.55 |
| snr_csf | 1.90 ± 0.35 | 1.88 ± 0.41 | 1.62 ± 0.28 | 2.00 ± 0.30 |
| snr_gm | 6.06 ± 1.01 | 6.16 ± 0.57 | 7.60 ± 0.73 | 5.52 ± 0.60 |
| snr_total | 6.16 ± 1.35 | 6.03 ± 0.90 | 8.06 ± 1.09 | 5.58 ± 0.93 |
| snr_wm | 10.52 ± 3.22 | 10.04 ± 2.17 | 14.97 ± 2.82 | 9.21 ± 2.20 |
| snrd_csf | -0.05 ± 0.92 | -0.63 ± 0.65 | -0.02 ± 1.26 | 0.14 ± 0.77 |
| snrd_gm | 0.71 ± 2.28 | -0.44 ± 1.13 | 1.35 ± 3.73 | 0.89 ± 1.76 |
| snrd_total | 0.70 ± 2.25 | -0.42 ± 1.17 | 1.30 ± 3.63 | 0.90 ± 1.76 |
| snrd_wm | 1.45 ± 3.62 | -0.20 ± 1.75 | 2.55 ± 5.96 | 1.66 ± 2.79 |
| summary_bg_k | 1.19 ± 3.15 | 0.95 ± 3.41 | 1.08 ± 3.16 | 1.31 ± 3.04 |
| summary_bg_mad | 226.94 ± 261.92 | 144.33 ± 303.34 | 72.10 ± 92.25 | 306.81 ± 252.63 |
| summary_bg_mean | 298.48 ± 322.63 | 245.98 ± 485.79 | 105.37 ± 128.77 | 380.54 ± 259.90 |
| summary_bg_median | 225.77 ± 274.46 | 206.56 ± 429.10 | 81.88 ± 103.54 | 280.00 ± 221.95 |
| summary_bg_n | 12742.22 ± 25443.44 | 1905.69 ± 8203.08 | 4702.95 ± 9812.76 | 19163.20 ± 30468.99 |
| summary_bg_p05 | 11.59 ± 51.78 | 24.98 ± 93.68 | 6.85 ± 26.43 | 8.51 ± 33.97 |
| summary_bg_p95 | 826.92 ± 831.47 | 586.29 ± 1116.30 | 287.23 ± 373.98 | 1088.89 ± 705.64 |
| summary_bg_stdv | 267.99 ± 268.52 | 178.40 ± 340.83 | 91.20 ± 120.37 | 357.55 ± 234.29 |
| summary_csf_k | 6.11 ± 22.41 | 12.91 ± 30.82 | 1.89 ± 7.31 | 5.14 ± 21.76 |
| summary_csf_mad | 93.64 ± 52.24 | 91.62 ± 12.76 | 88.65 ± 14.90 | 95.99 ± 66.62 |
| summary_csf_mean | 195.13 ± 45.64 | 199.45 ± 34.49 | 164.80 ± 22.69 | 203.65 ± 50.31 |
| summary_csf_median | 184.77 ± 47.91 | 186.48 ± 23.87 | 149.78 ± 28.29 | 195.74 ± 53.54 |
| summary_csf_n | 3469.30 ± 4434.52 | 2866.04 ± 2277.34 | 10157.86 ± 5881.93 | 1468.00 ± 935.11 |
| summary_csf_p05 | 54.34 ± 13.74 | 57.58 ± 11.28 | 41.29 ± 8.91 | 57.53 ± 13.29 |
| summary_csf_p95 | 369.52 ± 131.51 | 386.19 ± 190.16 | 334.93 ± 27.59 | 375.16 ± 125.48 |
| summary_csf_stdv | 101.30 ± 49.90 | 109.31 ± 66.04 | 92.80 ± 8.68 | 101.33 ± 50.98 |

| | | | | |
|---|---|---|---|---|
| summary_gm_k | 0.11 ± 0.23 | 0.04 ± 0.16 | 0.15 ± 0.39 | 0.12 ± 0.17 |
| summary_gm_mad | 97.67 ± 14.06 | 90.28 ± 7.11 | 80.11 ± 7.28 | 106.04 ± 10.29 |
| summary_gm_mean | 584.29 ± 30.34 | 550.31 ± 24.86 | 605.82 ± 22.86 | 588.98 ± 24.12 |
| summary_gm_median | 586.53 ± 31.35 | 552.29 ± 25.86 | 609.28 ± 23.57 | 590.90 ± 25.24 |
| summary_gm_n | 6233.90 ± 3146.11 | 8451.80 ± 2682.40 | 8068.69 ± 3485.69 | 4857.12 ± 2323.90 |
| summary_gm_p05 | 417.43 ± 37.57 | 398.93 ± 28.07 | 467.34 ± 27.60 | 407.36 ± 28.29 |
| summary_gm_p95 | 743.29 ± 38.28 | 694.34 ± 23.69 | 732.08 ± 22.10 | 764.00 ± 28.29 |
| summary_gm_stdv | 98.99 ± 14.40 | 90.12 ± 6.43 | 80.75 ± 6.48 | 108.10 ± 9.97 |
| summary_wm_k | 0.17 ± 0.36 | 0.08 ± 0.31 | 0.31 ± 0.24 | 0.15 ± 0.40 |
| summary_wm_mad | 101.97 ± 32.32 | 103.17 ± 24.57 | 66.47 ± 14.41 | 113.29 ± 30.50 |
| summary_wm_mean | 1001.03 ± 4.49 | 999.71 ± 4.65 | 999.86 ± 2.79 | 1001.88 ± 4.70 |
| summary_wm_median | 999.98 ± 0.04 | 999.99 ± 0.03 | 1000.00 ± 0.02 | 999.97 ± 0.04 |
| summary_wm_n | 128008.24 ± 21748.21 | 121130.17 ± 18390.11 | 135606.30 ± 20615.43 | 127885.67 ± 22393.33 |
| summary_wm_p05 | 832.27 ± 46.83 | 827.33 ± 36.05 | 885.23 ± 18.18 | 816.48 ± 44.00 |
| summary_wm_p95 | 1173.32 ± 50.92 | 1170.67 ± 38.09 | 1114.03 ± 28.33 | 1193.84 ± 44.71 |
| summary_wm_stdv | 103.49 ± 29.05 | 104.09 ± 21.49 | 69.31 ± 13.72 | 114.58 ± 26.17 |
| tpm_overlap_csf | 0.12 ± 0.02 | 0.12 ± 0.01 | 0.14 ± 0.01 | 0.11 ± 0.01 |
| tpm_overlap_gm | 0.37 ± 0.03 | 0.37 ± 0.03 | 0.40 ± 0.03 | 0.36 ± 0.03 |
| tpm_overlap_wm | 0.46 ± 0.03 | 0.46 ± 0.03 | 0.47 ± 0.03 | 0.46 ± 0.03 |
| wm2max | 0.39 ± 0.11 | 0.31 ± 0.06 | 0.58 ± 0.06 | 0.35 ± 0.07 |

**Supplementary Table 2** Spearman correlations (ρ) between artifact severity (ordinal scale 1–10) and four examples of image quality metrics (IQMs), from entropy, image contrast, inhomogeneity, and normative IQM categories, respectively. Positive values indicate that the IQM increases with higher artifact severity. EFC = entropy focus criterion, CNR = contrast-to-noise ratio, INU = intensity non-uniformity, TPM = tissue probability maps overlap.

| **artifact** | **EFC** | **CNR** | **INU** | **TPM** |
|---|---|---|---|---|
| **blurring** | 0.68 | 0.56 | -0.24 | 0.33 |
| **ghosting** | -0.15 | 0.05 | 0.35 | -0.31 |
| **motion** | -0.01 | 0.03 | 0.34 | -0.42 |
| **noise** | -0.71 | -0.21 | 0.50 | -0.14 |

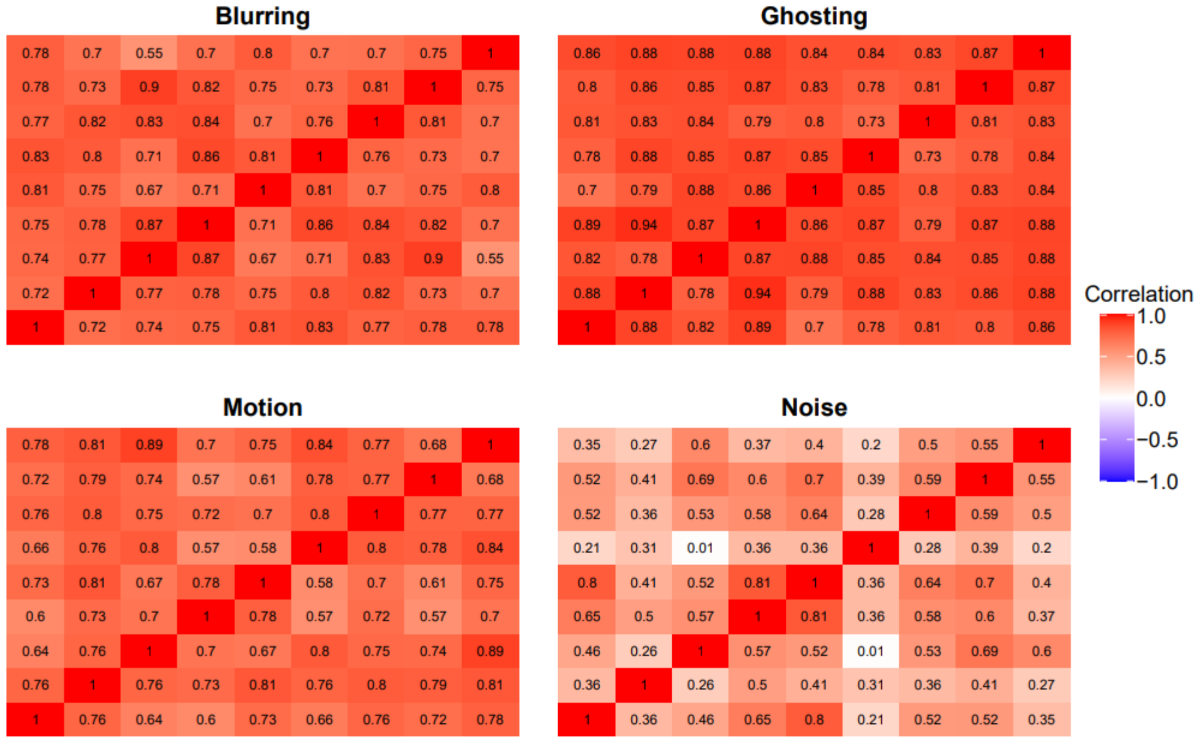


**Supplementary Figure 1.** Pairwise Spearman's rank correlation coefficients between raters for each artifact type (blur, ghosting, motion, and noise). Each heatmap shows the correlation between every pair of raters based on their visual QC ratings for a given artifact.

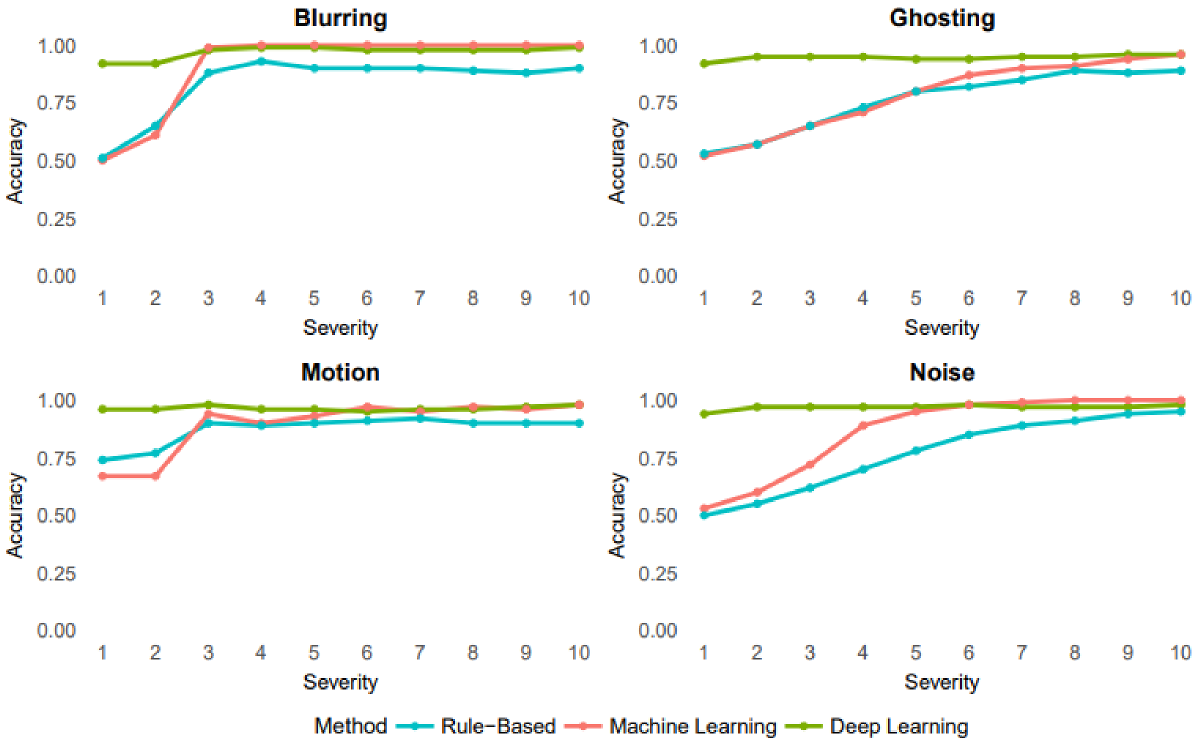


**Supplementary Figure 2**. Accuracy scores across artifact severity levels (1–10) for each artifact type (blur, ghosting, motion, and noise), comparing performance of rule-based (blue), machine learning-based (red), and deep learning-based (green) classification methods. Each panel presents results for a single artifact type.

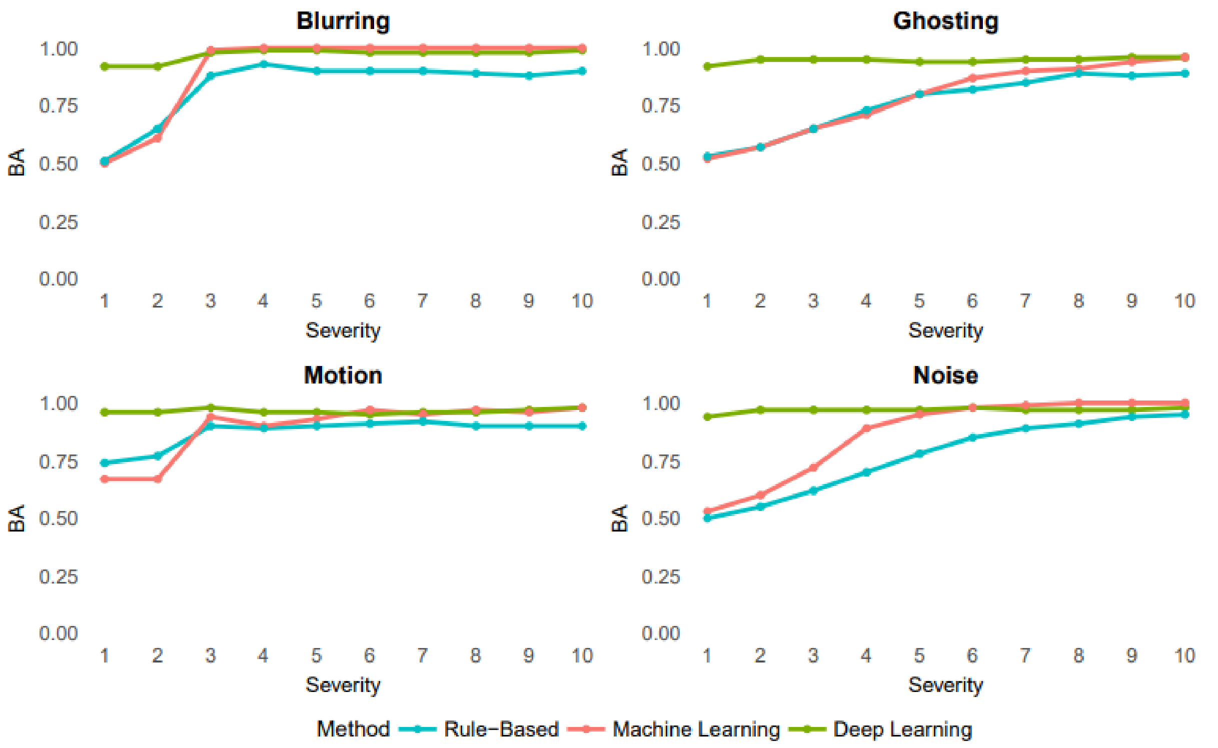


**Supplementary Figure 3.** Balanced Accuracy scores across artifact severity levels (1–10) for each artifact type (blur, ghosting, motion, and noise), comparing performance of rule-based (blue), machine learning-based (red), and deep learning-based (green) classification methods. Each panel presents results for a single artifact type. Lines represent the Balanced Accuracy ((sensitivity + specificity)/ 2) at each severity level.

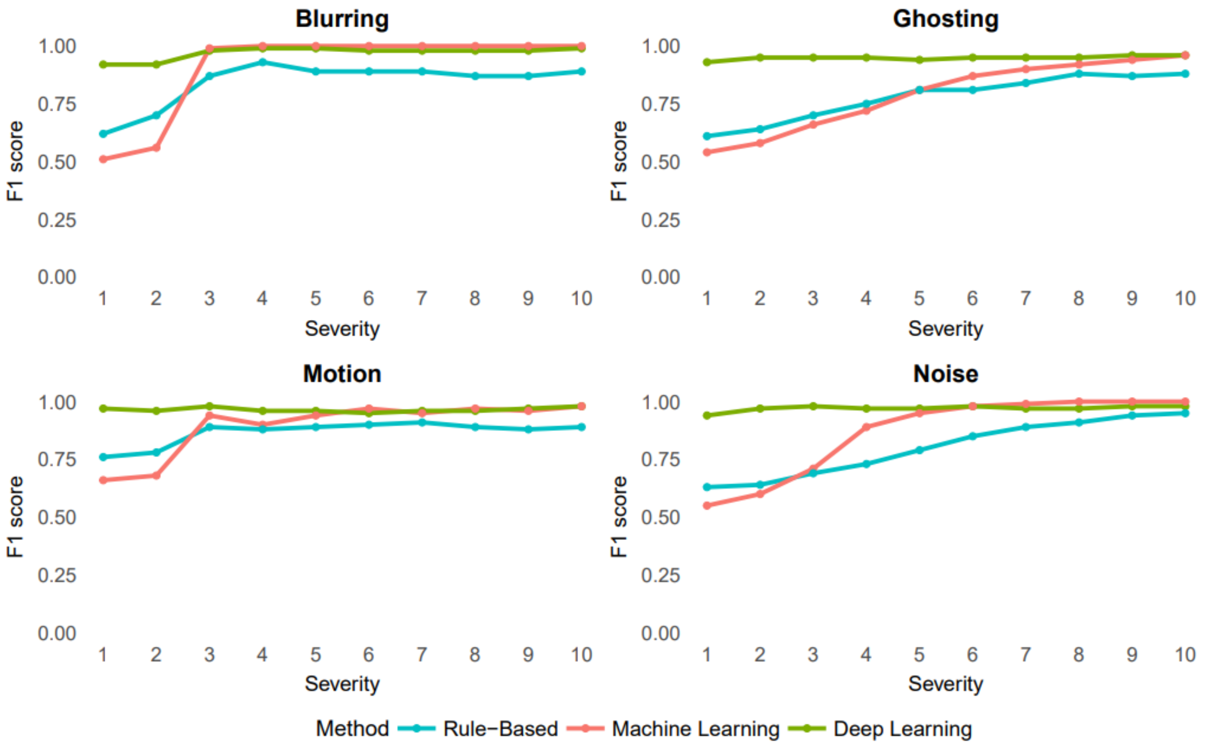


**Supplementary Figure 4**. F1 scores across artifact severity levels (1–10) for each artifact type (blur, ghosting, motion, and noise), comparing performance of rule-based (blue), machine learning-based (red), and deep learning-based (green) classification methods. Each panel presents results for a single artifact type.

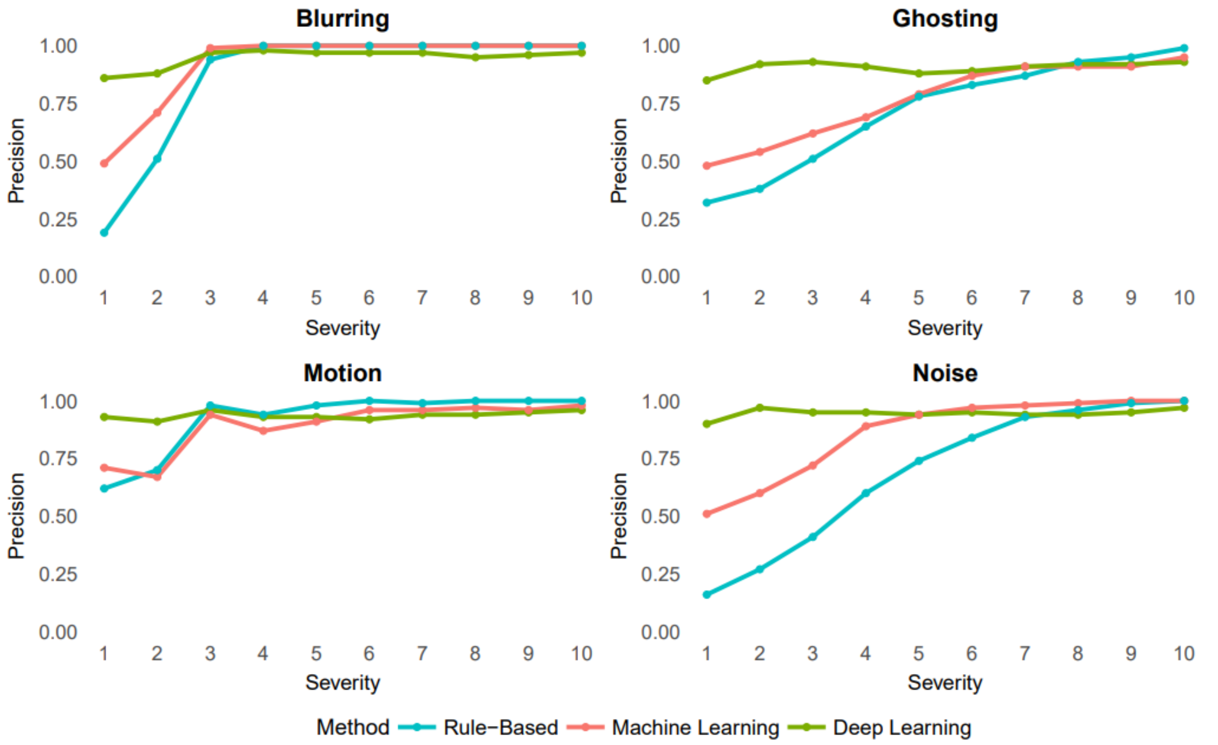


**Supplementary Figure 5**. Precision scores across artifact severity levels (1–10) for each artifact type (blur, ghosting, motion, and noise), comparing performance of rule-based (blue), machine learning-based (red), and deep learning-based (green) classification methods. Each panel presents results for a single artifact type.

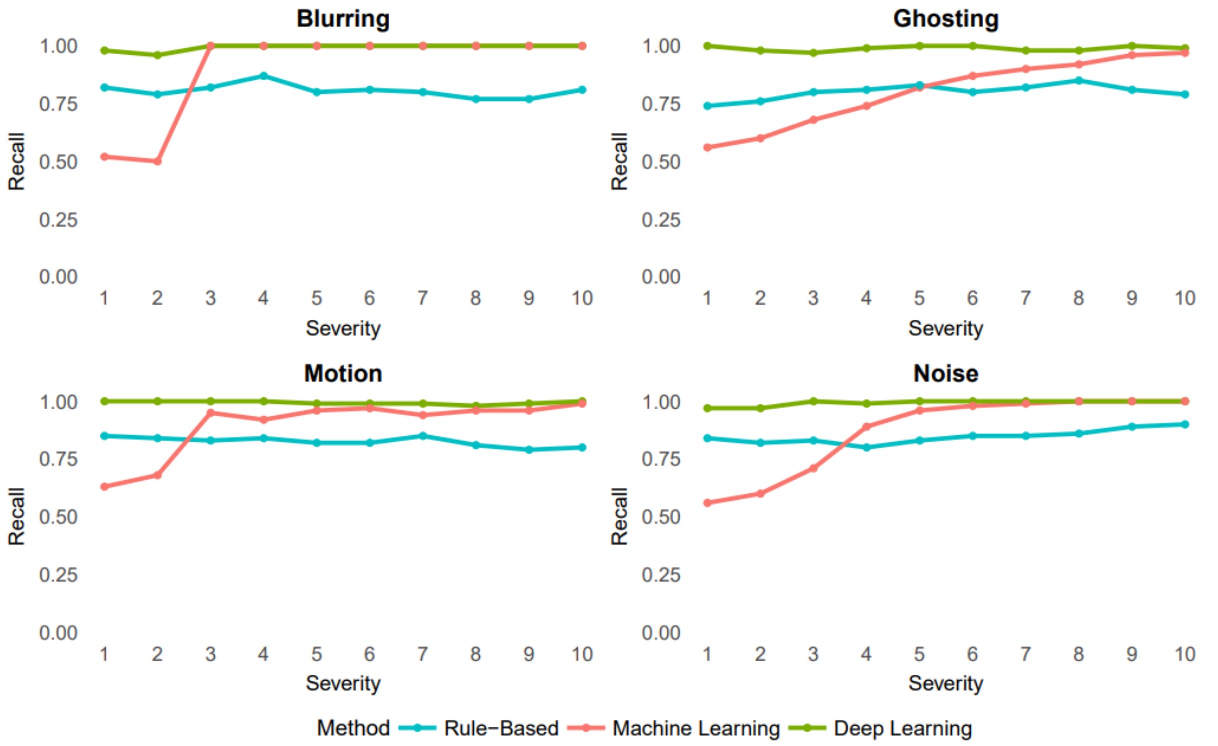


**Supplementary Figure 6**. Recall scores across artifact severity levels (1–10) for each artifact type (blur, ghosting, motion, and noise), comparing performance of rule-based (blue), machine learning-based (red), and deep learning-based (green) classification methods. Each panel presents results for a single artifact type.

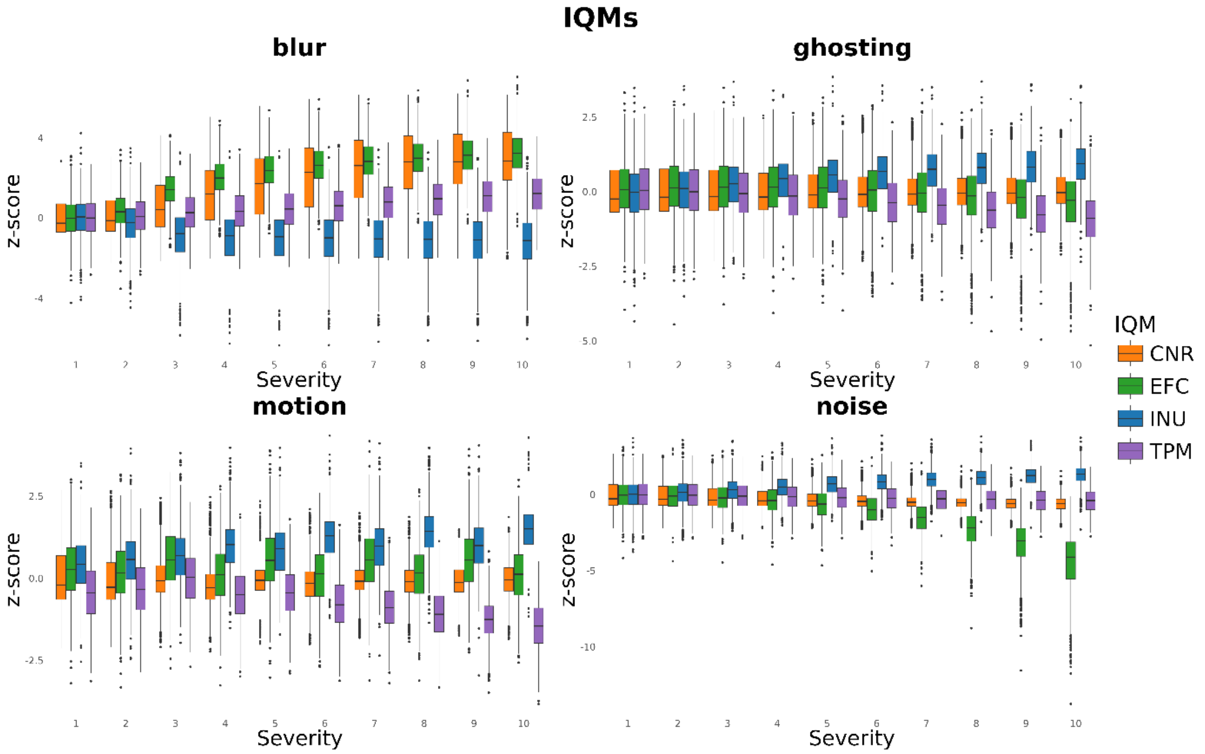


**Supplementary Figure 7.** Patterns of IQMs across artifact types and severity levels. Each panel displays z-scored values for four representative IQMs: contrast-to-noise ratio (CNR, orange), entropy focus criterion (EFC, green), median intensity non-uniformity (INU, blue), and tissue probability map overlap (TPM, purple) plotted as boxplots across severity levels.
